# Association between Resilience and Financial Toxicity: The Moderating Role of Age, Race and Socioeconomic Factors

**DOI:** 10.64898/2026.10.19.26365649

**Authors:** Yan Zhan, Rachel Wells, M. Tish Knobf, Liang Shan, Maryam Lustberg, Hsiao-Lan Wang

## Abstract

**PURPOSE:** Women with metastatic breast cancer (MBC) are at risk of developing financial toxicity due to prolonged treatment. Resilience has been identified as a key protective factor associated with better quality of life among cancer populations. However, it is unknown if resilience is associated with financial toxicity. We sought to examine the relationship between resilience and financial toxicity among women with MBC and explore whether socioeconomic factors moderate this association.

**PATIENTS AND METHODS:** Our sample included 209 women with MBC who completed self-reported questionnaires of the Resilience Scale Specific to Cancer (RS-SC) and the Comprehensive Score for Financial Toxicity (COST). Participants (n=2) with missing data were excluded from analysis. We conducted multiple linear regression to examine the association of resilience with financial toxicity. Interaction terms were used to test moderating effects of age (18-39, 40-64 and ≥65 years), race, education, health insurance, household income and employment status.

**RESULTS:** Of the sample, 56.5% were aged 40-64 years, 76.6% were White, and 79.9% had a college or master’s degree or above. Women reported moderately high resilience with mean RS-SC score of 34.49 (*SD* = 6.67), and moderate financial toxicity (mean COST score =21.01 [*SD* = 10.51]). Adjusting for covariates, higher resilience was linked to better financial well-being (b = 0.37, SE= 0.11, 95% CI [0.14, 0.59], *P* < 0.01). Age, education and health insurance moderated this relationship, with resilience more strongly associated with better financial well-being among those aged 40-64 years, with college/master’s degree or above and private insurance. Race, household income, and employment status did not moderate the association between resilience and financial toxicity.

**CONCLUSION:** Women with MBC who reported higher resilience are more likely to experience better financial well-being, but the strength of this association varied by socioeconomic factors. Further research is needed to confirm these findings. Our results have important implications for incorporating resilience into financial toxicity interventions and tailoring such interventions based on individual socioeconomic characteristics.

## Introduction

In the United States, the number of women living with metastatic breast cancer (MBC) has increased by 21% in the past decade (Mariotto et al., 2017), partly due to advances in new life-prolonging treatments, such as immunotherapy and targeted agent (Caswell-Jin et al., 2018). However, living longer with MBC was also accompanied by heightened costs for life-long cancer treatment, and use of palliative and supportive care (Sorensen et al.,2012). By 2030, the estimated overall annual cost for MBC in the United States will increase to $152.4 billion, which is an increase of 140% since 2015 (Gogate et al., 2021). A current systematic review reported that up to 27.3%-44.4% of women with breast cancer in high income countries developed financial toxicity (Ehsan et al., 2023), defined as the material hardship, psychological distress, and behavioral coping responses to the high cost of cancer care (Abrams at al., 2021; Carrera et al., 2018; Altice et al., 2017). Financial toxicity is a persistent life stressor contributing to worse mental health, adherence to treatment and quality of life (Arastu et al., 2020).

Studies in cancer populations have recognized resilience as an important protective factor that enables an individual patient to manage cancer-related stressors which has been associated with better quality of life and overall well-being (Aizpurua-Perez & Perez-Tejada, 2020; Ye et al., 2017, Seiler & Jenewein, 2019). Resilience is defined by the American Psychological Association as the process of successfully adapting to adversity or life challenges (American Psychological Association, 2020), and resilient individuals were often described as having the ability to “bounce back” from life stressors (American Psychological Association, 2020; Haase et al., 2017). Higher resilience in people with breast cancer was associated with better health-related quality of life and improved psychosocial outcomes, such as anxiety and depression (Zou et al., 2018; Mohlin et al., 2021; Matzka et al., 2016). However, how resilience operates in the context of financial toxicity for patients with advanced cancer and prolonged treatments is unknown. In addition, much of current literature on financial toxicity has predominately focused on identifying risk factors of financial hardship (Yabroff et al., 2018), whereas less attention has been paid to patient’s own resilience or ability to maintain financial well-being. To address this gap, it is critical to examine the association between resilience and financial toxicity.

Lower educational attainment, inadequate health insurance coverage (Yabroff et al., 2019), lower annual income, unemployment, and high out-of-pocket expenses (Smith et al., 2019; Sideris et al., 2025) have been identified as risk factors for financial toxicity. Among women with breast cancer, those who were single, younger, diagnosed with advanced breast cancer (Stage III and Stage IV or Metastatic stage) and receiving chemotherapy were more likely to experience financial toxicity than their counterparts (Celik et al., 2023). These socioeconomic factors may change how resilience impacts financial toxicity. In addition, racial/ethnic minority groups were also identified to be at higher risk of developing financial toxicity than individuals identified as non-Hispanic White (Chen et al., 2026; Pisu et al., 2015; Wheeler et al., 2018). Therefore, understanding how socioeconomic factors, age, and race moderate the association between resilience and financial toxicity, may help identify groups that may be most likely to benefit from resilience-enhancing strategies.

In this study, we aimed to examine if resilience is associated with financial toxicity and explored whether socioeconomic factors moderated this association among women with MBC. We hypothesized that higher resilience would be associated with better financial well-being, and this association may differ by age, race and socioeconomic factors: education level, health insurance, household income and employment status.

## Methods

### Research Design

We employed a descriptive cross-sectional study design with data collected from women living with MBC in the U.S. A detailed description of the study design and recruitment procedures has been reported in our previous study (Zhan et al., 2026). In brief, a total of 217 women who were diagnosed with MBC, currently receiving systemic cancer treatment, and had the ability to read and understand English were recruited from a comprehensive cancer center in Northeastern of the US and five MBC patient advocacy groups across the country from February to September 2024. We excluded women who were unable to provide informed consent to the study and who had a second cancer diagnosis at the same time. The study was approved by the Institutional Review Board of Yale University. All participants provided informed consent prior to participating in the study.

## Measures

### Resilience

A 10-item Resilience Scale Specific to Cancer (RS-SC-10) was used to measure resilience. Ye and colleagues (Ye et al., 2018) developed this scale to capture aspects of resilience relevant to patients with cancer, such as learning self-management skills and coping with fear of death. The example items included “I can adapt to changes in my surroundings when faced with cancer diagnosis” and “I can handle emotional distress”. Each item was rated on a 5-Likert scale ranging from 1= “Never” to 5= “Always”. The total scores range from 10 to 50, with higher scores indicating higher levels of resilience. The RS-SC-10 has been validated among patients with breast cancer in the United States, with a Cronbach’s α of 0.83 in the present study.

### Demographics and socioeconomic variables

Demographic information and socioeconomic data were collected by a self-reported questionnaire, including age, race, marital status, household income, employment status, education level, and health insurance type. Participants aged 18-39 years were defined as young adults (Fidler et al., 2017). Middle-aged adults and older adults were defined as 40-64 years and ≥ 65 years, respectively based on age eligibility for Medicare insurance coverage (Yabroff et al., 2019) and United States Census guidelines for the adult categories (Lindemann et al., 2017). Clinical information, including time since MBC diagnosis and cancer treatment types (i.e., chemotherapy, hormone therapy, targeted therapy and immunotherapy), was also collected.

### Financial toxicity

We used the Comprehensive Score for Financial Toxicity (COST) (version 2) to measure financial toxicity. The COST is a validated patient-reported measure developed by de Souza and colleagues to describe financial distress among patients with advanced cancer, with three specific domains: financial resources, financial burden and psychological distress (de Souza et al., 2017). The scale consists of 12 items and was rated on a five-point Likert scale from 0 = not at all to 4= very much. The final question was a summary item and was not required to calculate the total score. The remaining 11 items were summed to generate a total score ranging from 0 to 44, with higher COST scores indicating lower financial toxicity (better financial well-being), or vice versa. The Cronbach’s α was 0.90 for the COST in our study, indicating an excellent internal consistency.

### Data Analysis

We used R version 4.5.3 to conduct all statistical analysis. Categorical variables were dichotomized to enhance interpretability. Descriptive statistics were used to summarize sample characteristics. We calculated frequencies and percentages for categorical variables and mean, and standard deviation were reported for continuous variables. Analysis of variance (ANOVA) was employed to test the difference in financial toxicity among different groups. Pearson and spearman correlation coefficients were used to examine the association between continuous variables. Then, we conducted hierarchical multiple linear regression to examine the associations between resilience and financial toxicity, adjusting for demographics (age, race, marital status), socioeconomic variables (household income, employment status, educational level, and health insurance), and clinical factors (time since MBC diagnosis, and cancer treatment) sequentially. The variance inflation factor (VIF) values for variables ranged from 1.06 to 1.29, indicating no multicollinearity among variables. Finally, we performed moderation analyses using interaction terms between resilience, age, race and socioeconomic factors to examine whether the association between resilience and financial toxicity differed across subgroups. A two-sided P value of < 0.05 was set for statistical significance in this study.

## Results

### Characteristics of participants

A total of 217 women with MBC participated in the study. After excluding eight invalid questionnaires, the final analytic sample was 209. As shown in Table 1, participants in this study were predominantly White (76.6%), married (70.3%), and the majority had a college degree or master’s degree or above (79.9%). More than half of the participants were middled aged women (56.5%). While all had health insurance coverage, with public health insurance was the primary source of coverage for 57.9% of the participants. The median length of time since the first MBC diagnosis was 3 years (mean ± SD, 4.35 ± 3.98) with slightly more than half of the participants reported receiving hormone therapy (53.6%) or targeted therapy (53.1%).

**Table 1.**
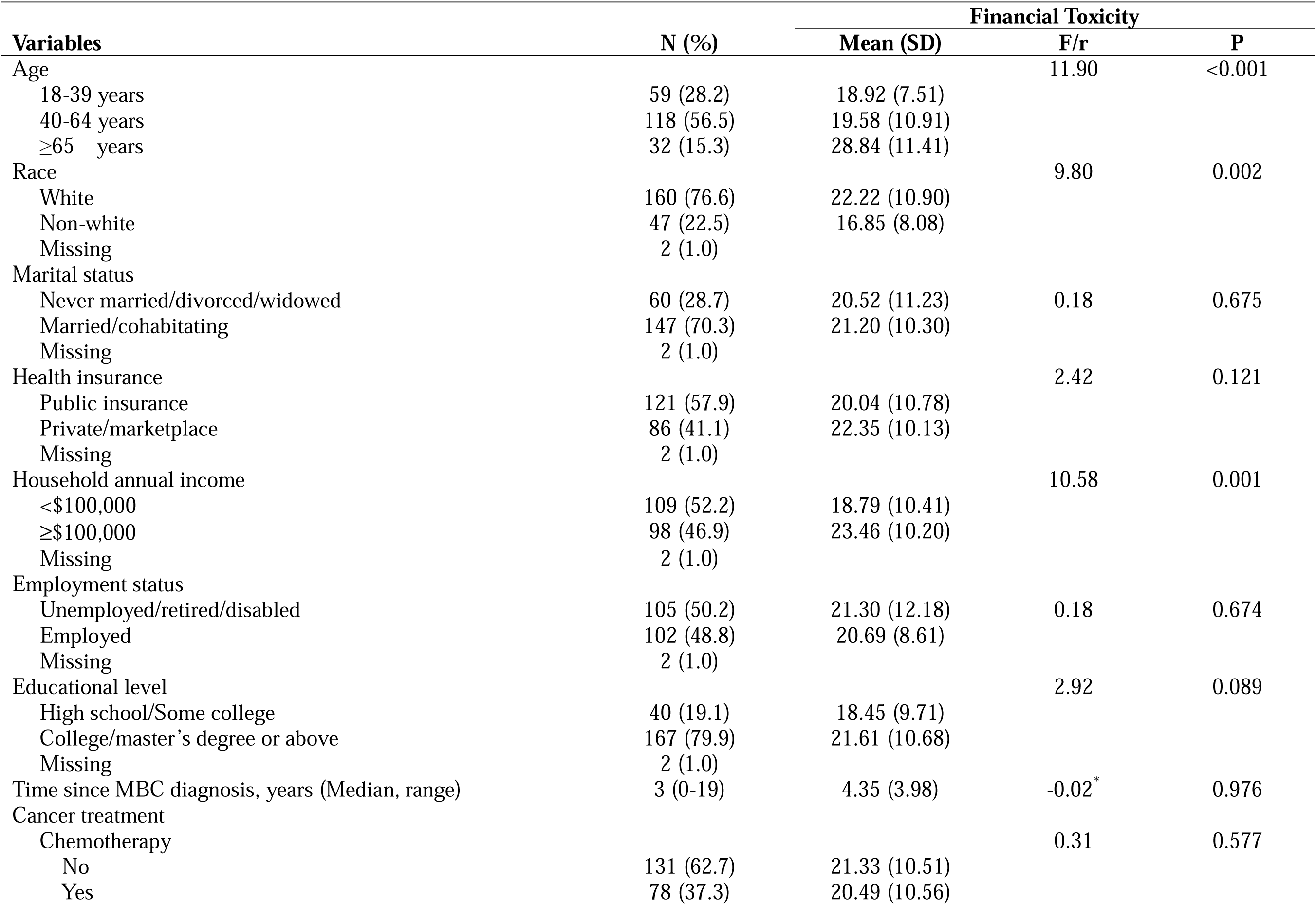
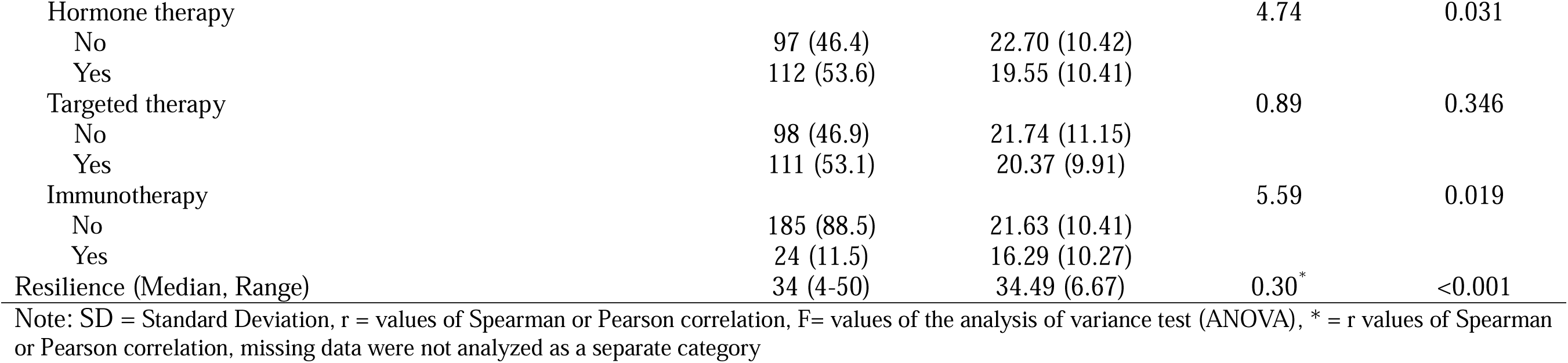
Characteristics of Women with Metastatic Breast Cancer (N=209)

Significant differences in mean COST scores were identified by age group, race and annual household income. COST scores differed significantly across age groups (P < 0.001) with participants aged ≥65 years reporting the highest mean score of 28.84 (SD ± 11.41), followed by those aged 40-64 years (mean ± SD, 19.58 ± 10.91) and 18-39 years (mean ± SD 18.92 ± 7.51). Women who identified as White showed higher mean COST scores than those identified as non-White (mean ± SD, 22.22 ± 10.90 vs. 16.85 ± 8.08). Participants with an annual household income of $100,000 or greater reported better financial well-being than those with an income below $100,000 (mean ± SD, 23.46 ± 10.20 vs. 18.79 ± 10.41). The mean score of resilience was 34.49 (SD = 6.67), indicating overall moderate level of resilience and resilience scores were positively correlated with the COST scores (r = 0.30, P < 0.001).

### Multiple linear regression models

Hierarchical multiple linear regression model that examined factors associated with COST scores are detailed in Table 2. First, we conducted an unadjusted model to include only resilience as a single predictor of COST scores. Then, we controlled for demographic characteristics, socioeconomic factors and clinical characteristics sequentially in each model. The results showed that resilience was positively associated with higher COST scores (b = 0.37, SE= 0.11, 95% CI [0.14, 0.59], *P* < 0.01), adjusting for demographic, socioeconomic, and clinical related factors.

**Table 2.**
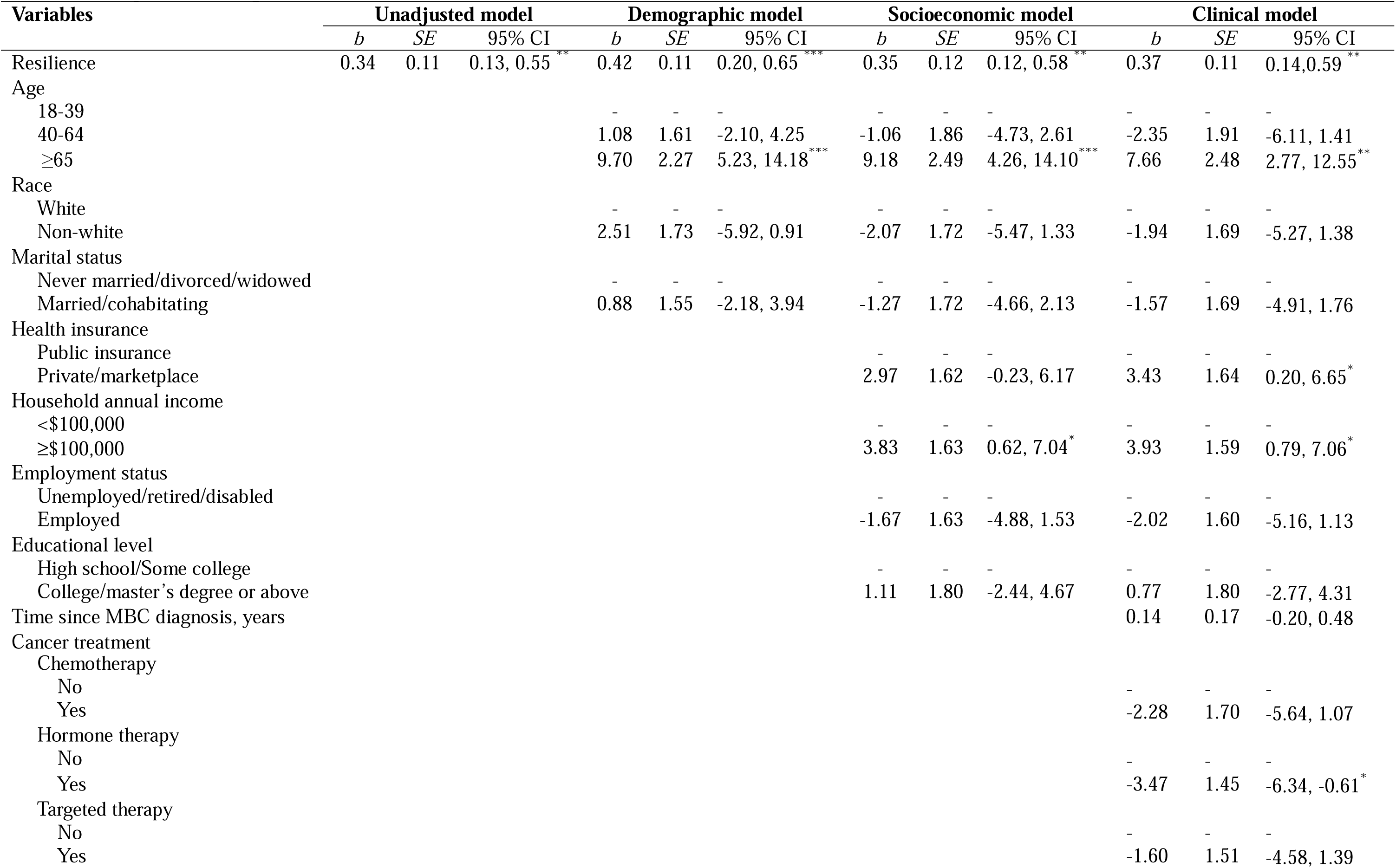
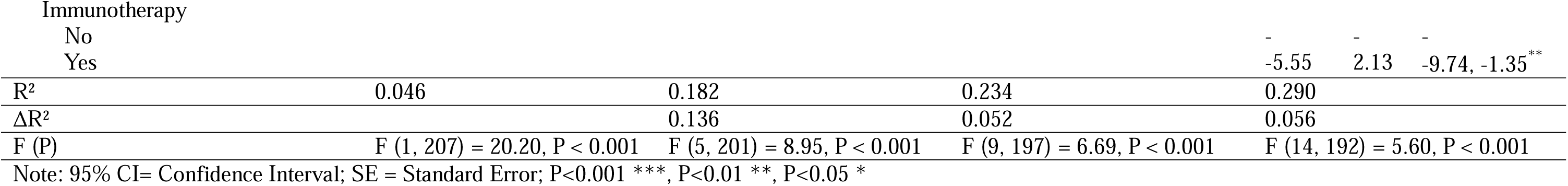
Multiple Linear Regression Model.

Among demographic factors, older age (age ≥ 65 years) was the only factor that was associated with lower financial toxicity (higher COST scores). When controlling for socioeconomic factors, annual household income was associated with higher COST scores (b = 3.83, SE = 1.63, 95% CI [0.62, 7.04], p < 0.05), whereas other economic factors including health insurance, employment status and educational level were not associated with COST scores. In the clinical model, receiving hormone therapy and immunotherapy were both independently associated with lower COST scores. In contrast, private or marketplace insurance was associated with higher COST scores after fully adjusting for clinical factors.

### Moderation analyses

We tested interaction terms between resilience and each potential moderator: age, race, and four socioeconomic factors (education, health insurance, household income, and employment status), in separate linear regression models. Results of the moderation analyses were presented in Table 3.

**Table 3.** Interaction models between resilience and age, race and socioeconomic variables.

| **Variables** | ***b*** | ***SE*** | **95% CI** | **$\beta$** | ***P*** |
| --- | --- | --- | --- | --- | --- |
| **1. Age** |  |  |  |  |  |
| Resilience | -0.02 | 0.19 | -0.39, 0.36 | -0.01 | 0.929 |
| Age (ref: 18-39) | - | - | - | - | - |
| 40-64 | -27.34 | 8.92 | -44.94, -9.74 | -1.29 | 0.002 |
| ≥65 | -0.63 | 11.69 | -23.68, 22.42 | -0.02 | 0.957 |
| *Interaction terms* |  |  |  |  |  |
| Resilience × Age | - | - | - | - | 0.014 |
| Resilience × 40-64 | 0.71 | 0.25 | 0.22, 1.19 | 1.19 | 0.005 |
| Resilience × ≥65 | 0.22 | 0.33 | -0.42, 0.87 | 0.27 | 0.496 |
| **2. Race** |  |  |  |  |  |
| Resilience | 0.42 | 0.13 | 0.17, 0.67 | 0.24 | 0.001 |
| Race (ref: White) | - | - | - | - | - |
| Non-white | 6.22 | 9.70 | -12.91, 25.34 | 0.25 | 0.522 |
| *Interaction terms* |  |  |  |  |  |
| Resilience × Non-white | -0.24 | 0.28 | -0.80, 0.31 | -0.32 | 0.394 |
| **3. Education** |  |  |  |  |  |
| Resilience | -0.05 | 0.19 | -0.42, 0.32 | -0.03 | 0.790 |
| Education (ref: High school/Some college) | - | - | - | - | - |
| College/master's degree or above | -21.02 | 8.15 | -37.09, -4.95 | -0.79 | 0.011 |
| *Interaction terms* |  |  |  |  |  |
| Resilience × College/master's degree or above | 0.63 | 0.23 | 0.18, 1.09 | 0.88 | 0.007 |
| **4. Health insurance** |  |  |  |  |  |
| Resilience | 0.17 | 0.14 | -0.11, 0.45 | 0.10 | 0.228 |
| Health insurance (ref: Public) | - | - | - | - | - |
| Private/marketplace | -15.80 | 8.27 | -32.11, 0.51 | -0.74 | 0.058 |
| *Interaction terms* |  |  |  |  |  |
| Resilience × Private/marketplace | 0.54 | 0.23 | 0.09, 1.00 | 0.93 | 0.019 |
| **5. Household income** |  |  |  |  |  |
| Resilience | 0.19 | 0.15 | -0.09, 0.48 | 0.11 | 0.187 |
| Household income (ref: <\$100,000) | - | - | - | - | - |
| ≥\$100,000 | -11.58 | 8.20 | -27.75, 4.58 | -0.55 | 0.159 |
| *Interaction terms* |  |  |  |  |  |
| Resilience × ≥\$100,000 | 0.44 | 0.23 | -0.01, 0.89 | 0.77 | 0.055 |
| **6. Employment status** |  |  |  |  |  |
| Resilience | 0.43 | 0.15 | 0.13, 0.73 | 0.24 | 0.005 |
| Employment status (ref: Unemployed/retired/disabled) | - | - | - | - | - |
| Employed | 3.20 | 8.39 | -13.35, 19.75 | 0.15 | 0.704 |
| *Interaction terms* |  |  |  |  |  |
| Resilience × Employed | -0.15 | 0.23 | -0.60, 0.31 | -0.26 | 0.527 |

Note: 95% CI= Confidence Interval; SE = Standard Error; All models adjusted for all other covariates with the moderator of interest included as a main effect along with its interaction term with resilience

### Age

As shown in Table 3, age significantly moderated the association between resilience and COST scores (resilience × age, P = 0.014). Compared with participants aged 18-39 years (reference group), positive association between resilience and COST scores was only significant among those aged 40-64 years (b = 0.71, SE = 0.25, 95% CI [0.22, 1.19], p = 0.005). Participants aged ≥65 years did not differ from the reference group (b = 0.22, SE = 0.33, 95% CI [−0.42, 0.87], p = .496).

### Education

Education moderated the association between resilience and COST scores (b = 0.63, SE = 0.23, 95% CI [0.18, 1.09], p = 0.007). Higher resilience levels were significantly associated with higher COST scores (lower financial toxicity) among participants with a master’s degree or above. However, among participants with lower education attainment (i.e., high school/some college), resilience was not associated with COST scores.

### Health insurance

Health insurance type was also identified as a strong moderator of resilience and COST scores. The interaction between resilience and health insurance type was significant (b = 0.54, SE = 0.23, 95% CI [0.09, 1.00], p = 0.019), indicating that the positive association between higher resilience and higher COST scores was stronger among participants with private or marketplace insurance than among those with public insurance (i.e., Medicaid and Medicare). Nevertheless, this association was not observed among participants with public insurance.

We did not observe any interaction effects between resilience and race, household income, or employment status (Table 3).

## Discussion

This study is among the first to examine the association between resilience and financial toxicity among women with MBC and identify important demographic and socioeconomic factors that moderate this relationship. Findings from this study can be used to guide the development of age appropriate and socioeconomically tailored interventions that promote resilience in navigating financial toxicity among women with MBC and other cancer populations.

Consistent with our hypotheses, the findings of this study identified that resilience was strongly associated with lower financial toxicity (better financial well-being), even after controlling for demographic, socioeconomic and clinical variables. When a woman is diagnosed with MBC and begin cancer treatment, she must adapt to not only the physical and emotional burden of cancer but also accumulating financial stressors including direct medical costs of co-payment and out-of-pocket, as well as indirect costs related to transportation, lodging, and employment disruption (Lee et al., 2023; Ko et al., 2025; Marshall et al., 2025). The American Psychological Association suggested three key factors that determine successful adaptation to stressors, including how individuals view and engage with their circumstances, the availability and quality of social resources, and skills associated with positive adaptation (American Psychological Association, 2022). How an individual woman appraises their circumstances and engages in managing financial stressors may directly influence their financial and health outcomes. Women with higher resilience may be more proactively engaged in positive coping strategies, for example, communicating financial concerns with the oncology clinician team (Lee 2023), negotiating with the insurance company (Jones et al., 2024) and seeking social support (Gharzai et al., 2021) and financial assistance (Prabandari et al., 2022; Enzler et al., 2019), all of which contribute to better financial well-being. Lack of resilience, on the other hand, may be associated with fewer financial resources, lower engagement in financial management behaviors and greater maladaptive coping and emotional distress. However, the construct of resilience in the context of financial toxicity was still unclear and underexplored. Future qualitative study or mixed-methods research should be warranted to further explore specific resilience-based tasks through women’s experience, as well as the necessary resources that facilitate this process.

The association between resilience and financial toxicity varied across age groups. Aligned with previous studies (Li et al., 2025; Corrigan et al., 2022), women aged 18-39 and 40-64 years in our study had approximately one-third lower mean COST scores than those aged ≥65 years (18.92 and 19.58 vs. 28.84), indicating a greater financial toxicity level among younger and middle-aged women than the older group. It is interesting to note that the positive association between higher resilience and lower financial toxicity was only found in the middle-aged women. We posited that middle-aged women may have already established social, occupational and health resources, as well as gained life experience that enables them to cope financial stress, even when confronting employment disruption and medical debt incurred by MBC and cancer treatment. However, younger women, who were at an early stage of their life course, may have fewer financial reserves, lower financial literacy, and limited access to resources needed to mitigate financial toxicity compared with middle- and older-aged women (Corrigan et al., 2022). Therefore, even if resilience was strengthened for younger age group, it may not improve their financial toxicity when essential financial, social and supportive resources were unavailable to them. This finding highlighted the need to integrate multi-levels of resources or develop multi-component interventions that promote women’s resilience and financial well-being.

Among women with a college or master’s degree, higher resilience was associated with lower financial toxicity, but this association was insignificant among women with a lower educational level. One plausible explanation is that high education attainment can be related to higher insurance literacy and exposure to financial information that enable women to make reasonable financial and treatment decisions. Prior studies have shown strong links between low financial/insurance literacy levels and financial hardship (Lee et al., 2023; Khera et al., 2022). For well-educated women, resilience-promoting interventions may help improve self-efficacy and motivate them to access and use available resources to address financial toxicity. However, enhancing resilience alone may not be sufficient to alleviate financial toxicity among women who only attained high school of some college. While an individual may be psychologically adaptive, optimistic and hopeful, but those factors could not compensate for their limited access to financial and health resources. Thus, improving health related financial literacy and providing necessary resources may be important strategies to promote resilience in coping with financial toxicity among women with lower education levels.

Despite that financial toxicity scores as measured by COST did not differ significantly between women with private/marketplace insurance and those with public insurance, the moderation analysis indicated that resilience was only positively associated with better financial toxicity among women with private/marketplace insurance. A prior study found that patients with public insurance (Medicaid or Medicare) have greater risk of financial hardship than patients who have private insurance (OR, 1.95, P < .0001) and having public health insurance may also be associated with lower savings and assets, which contributes to financial vulnerability and decrease resources for resilience (Banegas et al., 2016). As noted, neither household annual income nor employment status moderated the association between resilience and financial toxicity. This result was contrary to our expectation, as these two factors are important indicators of financial toxicity and usually associated with health insurance status. However, income and employment may not directly capture the entire experience. There is evidence that even insured patients with private coverage may also experience financial burden when deductibles and out-of-pocket costs were high (Fu et al., 2021; Yabroff et al., 2024). Future work should focus on unpacking the interaction between income, employment and insurance and their collective effects on resilience and financial toxicity.

## Implications

Our results have important implications for clinicians who engaged with cancer patients and their families to address financial toxicity. Given the association between resilience and low financial toxicity, resilience-enhancing interventions may be initiated early in the first six months of cancer trajectory when many patients begin to face financial toxicity (Bell-Brown et al., 2024). This intervention may focus on assessing and promoting women’s resources for resilience and positive coping skills, such as managing cost-related distress and uncertainty, adjusting financial plans, and navigating resources and support from community and the broader health/social system. For women with low educational level and income, resilience-enhancing interventions may need to be tailored to improve their ability to cope with limited economic resources. However, cultivating resilience alone may be insufficient for preparing women well in the face of persistent financial challenges. Future resilience interventions can be integrated or combined with existing financial navigation or insurance counseling services to comprehensively address financial toxicity.

## Limitations

This study has several limitations to be considered. First, the cross-sectional design limits our capability to draw causal conclusions about the relationship between resilience and financial toxicity. Future longitudinal studies will be needed to examine whether resilience predicts changes in financial toxicity over time and if the strength of this association varies across the cancer trajectory. Second, financial toxicity was assessed using the COST measure, which only captured perceived financial well-being rather than objective financial burden and material hardship. Future studies should use objective indicators, such as out-of-pocket costs (Smith et al., 2019), medical debt, employment disruption (Hueniken et al., 2020), insurance details, and use of financial assistance (Yabroff et al., 2018), to better understand how subjective and objective financial toxicity may be associated with resilience. Third, socioeconomic variables such as income, employment, and insurance were measured using broad categories. For example, the category of public health insurance included different types of plans, such as Medicare and Medicaid, which may vary substantially in eligibility, benefits, cost-sharing, and supplemental coverage. This limitation can be addressed by using more granular measures of socioeconomic status and insurance coverage to better capture nuanced differences across patient groups. In addition, our sample consisted of highly educated, middle-class participants, which may limit the generalizability of the findings to other MBC groups, particularly those from socioeconomically disadvantaged communities who experience the greatest financial toxicity. Finally, some subgroup analyses may have been limited by small sample size. For example, the lower educational attainment group included only 40 participants, which may have reduced statistical power to detect moderation effects.

## Conclusion

Resilience was positively associated with better perceived financial well-being among our sample of 209 women with MBC, but this association varied by age, education level and health insurance status. Critical next steps in research include validating these results in other cancer populations and defining the construct of resilience within the context of financial toxicity. Future intervention development may focus on cultivating positive adaptation skills and increase access to supportive resources that optimize patients’ coping behaviors and financial well-being.

## Data Availability

All data produced in the present study are available upon reasonable request to the authors

